# Histopathological Image Analysis for Oral Squamous Cell Carcinoma classification using concatenated deep learning models

**DOI:** 10.1101/2021.05.06.21256741

**Authors:** Ibrar Amin, Hina Zamir, Faisal F. Khan

## Abstract

Oral squamous cell carcinoma (OSCC) is a subset of head and neck squamous cell carcinoma (HNSCC), the 7th most common cancer worldwide, and accounts for more than 90% of oral malignancies. Early detection of OSCC is essential for effective treatment and reducing the mortality rate. However, the gold standard method of microscopy-based histopathological investigation is often challenging, time-consuming and relies on human expertise. Automated analysis of oral biopsy images can aid the histopathologists in performing a rapid and arguably more accurate diagnosis of OSCC. In this study, we present deep learning (DL) based automated classification of 290 normal and 934 cancerous oral histopathological images published by Tabassum et al (Data in Brief, 2020). We utilized transfer learning approach by adapting three pre-trained DL models to OSCC detection. VGG16, InceptionV3, and Resnet50 were fine-tuned individually and then used in concatenation as feature extractors. The concatenated model outperformed the individual models and achieved 96.66% accuracy (95.16% precision, 98.33% recall, and 95.00% specificity) compared to 89.16% (VGG16), 94.16% (InceptionV3) and 90.83% (ResNet50). These results demonstrate that the concatenated model can effectively replace the use of a single DL architecture.

## 1. Introduction

Oral squamous cell carcinoma (OSCC), is a heterogeneous group of cancer arising from the mucosal lining of the oral cavity [1] accounting for more than 90% of oral malignancies [2]. It is a subset of head and neck squamous cell carcinoma (HNSCC), the 7th most common cancer worldwide [3]. According to World Health Organization, the estimated number of new cases each year is 657,000 with more than 330,000 deaths worldwide. The South Asian countries were observed to have significantly higher incidence rates of OSCC. Among them, India has the largest number of cases globally (one–third), while in Pakistan, it is the first and second most prevalent cancer in males and females, respectively [4]. The risk factors include use of alcohol and tobacco, poor oral hygiene, exposure to human papillomavirus (HPV), genetic background, lifestyle, ethnicity, and geographical location.

The early diagnosis of OSCC is crucial for effective treatment, improvement in survival chances, and reducing morbidity and death rates [5]. The OSCC has a poor prognosis with a 50% overall survival rate [6]. Currently, the gold standard for diagnosis of OSCC is microscopy-based histopathological investigation of tissue biopsies [7]. This diagnostic pathology method relies on the interpretation of histopathologists, which is usually slow and prone to error, thus limiting its clinical utility [8]. It is, therefore, essential to develop efficient diagnostic tools that can assist the pathologists in the analysis and diagnosis of OSCC.

Recently, there has been an expanding corpus of research on improving medical diagnosis using artificial intelligence (AI). The increased use of diagnostic imaging has enabled researchers to investigate AI applications in analyzing medical images. In particular, one AI technique, i.e., Deep Learning (DL), has shown significant successes in addressing several different medical image analysis problems[9], particularly in the diagnosis of cancer in pathological images [10]. Based on DL, Computer-Aided Diagnosis (CAD) systems have been proposed and developed on substantial scales for various cancer types such as breast cancer [11], lung cancer [12], prostate cancer [13], etc. However, for the diagnosis of OSCC from pathological images, the literature reveals that DL has been scarcely adopted. In a study to detect keratin pearls in oral histopathology images, Dev *et al* used Convolutional Neural Network (CNN) and Random Forest. The CNN model achieved 98.05% accuracy for keratin region segmentation, while the Random Forest detected keratin pearls with 96.88% accuracy [14]. Similarly, Das *et al* utilized DL to classify oral biopsy images into multiple classes as per Broder’s histological grading system. Furthermore, CNN was proposed that showed a classification accuracy of 97.5% [15]. Jonathan *et al* applied Active Learning (AL) and Random Learning (RL) through CNN for the classification of oral cancer tissue into seven classes (stroma, lymphocytes, tumor, mucosa, keratin pearls, blood, and adipose). It was found that the accuracy achieved by the AL surpassed that of RL by 3.26% [16]. Moreover, Francesco *et al* performed pixel-wise segmentation of oral lesion whole slide images (WSI) into three classes (carcinoma, non-carcinoma, and non-tissue) using different DL architectures, such as U-Net, SegNet, U-Net with VGG16 encoder, and U-Net with ResNet50 encoder. It was shown that a deeper network, such as U-Net modified with ResNet50 as encoder, had better accuracy than the original U-Net [17]. Recently Rutwik *et al* performed binary classification on oral pathology images and achieved an accuracy of 91.13% using ResNet [18].

In this paper, we improved the classification of OSCC histopathological images into normal and cancerous classes. This improvement is achieved by utilizing the concept of transfer learning through pre-trained CNN models (VGG16, ResNet50, and InceptionV3). The models were used both as individually fine-tuned classifiers and in a combination as feature extractors. We also provided a detailed analysis of models’ performance using various metrics to report the best network for OSCC detection.

## 2. Data and Methodologies

### 2.1. Experimental Setup

Experiments in this study were conducted on an HP Z8 workstation with NVIDIA P2000 GPU and 64 GB RAM. The network architectures were implemented using Python’s Keras library with Tensorflow backend.

### 2.2. Dataset

The OSCC dataset is publicly available and was published by Tabassum *et al* [19]. It is composed of 1224 oral histopathological images (290 non-cancerous and 934 cancerous) from 230 patients. Figure 1 shows instances of both the classes from the dataset. The images were captured at two different magnifications (100x and 400x) from Hematoxylin and Eosin (H&E) stained tissue slides using Leica ICC50 HD microscope. 89 images with the normal epithelium and 439 images of OSCC were in 100x magnification, while 201 normal and 495 OSCC images were in 400x magnification (Figure 2).

**Figure 1:**
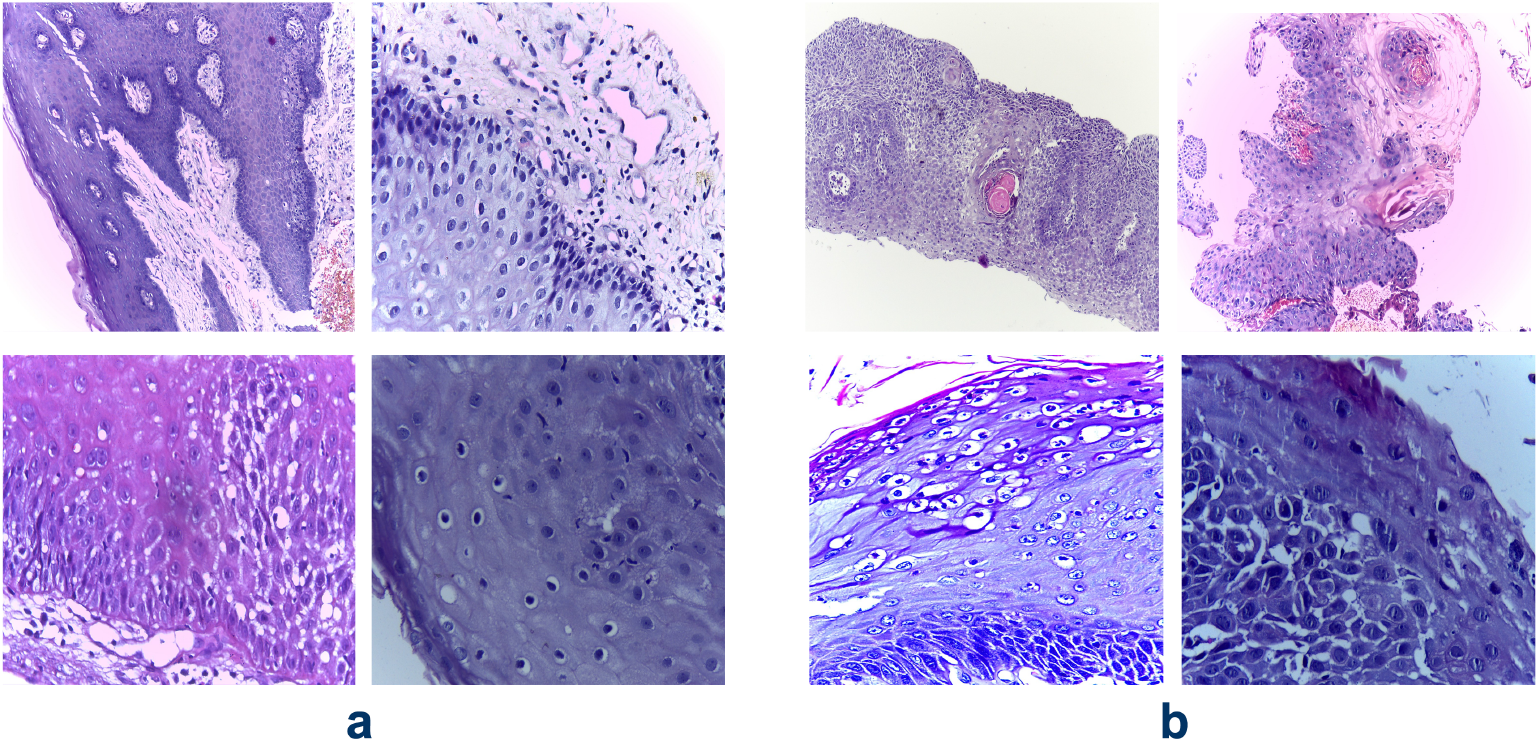
Vignettes of H&E stained oral biopsy images from the OSCC dataset by Tabassum *et al* [19] capturing normal epithelium (a) and cancerous epithelium(b)

**Figure 2:**
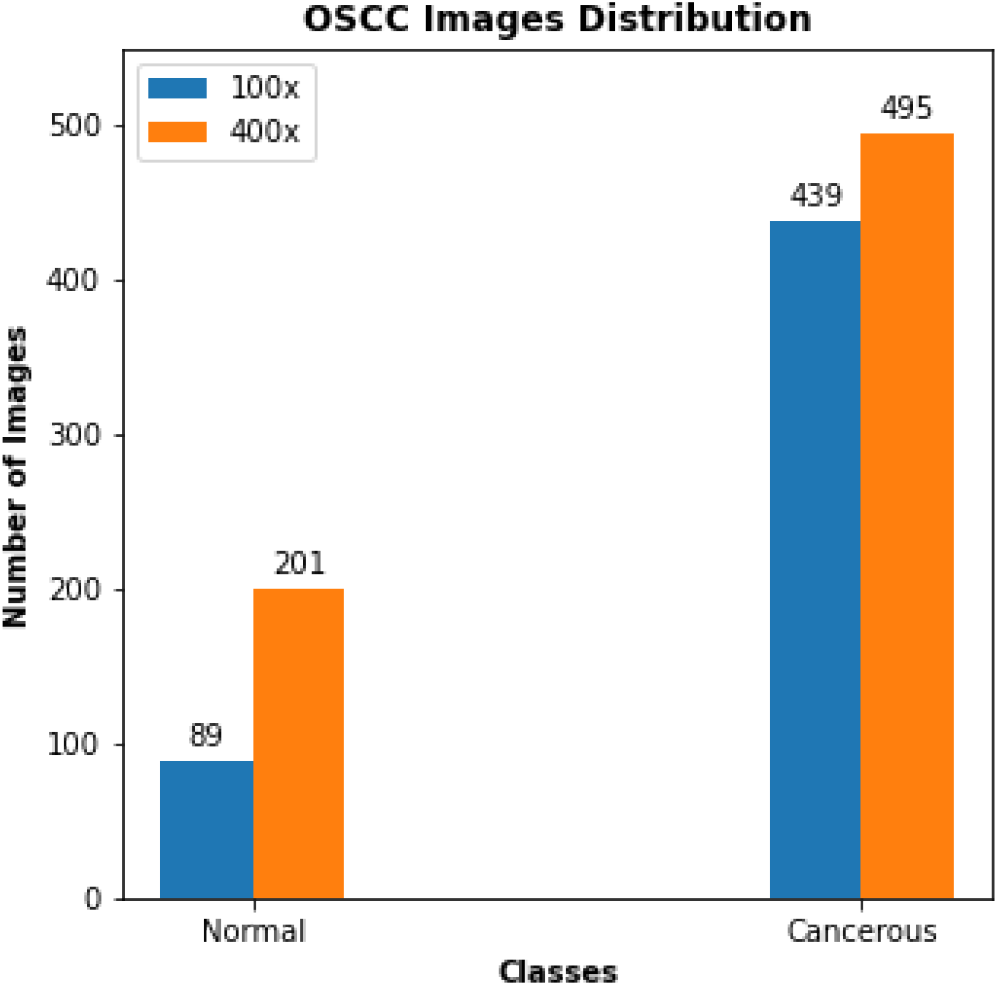
Distribution of images in the OSCC dataset.The Normal and Cancerous class contains 290 and 934 images, respectively. The images are in two different magnifications — 100x (89 Normal, 439 Cancerous) and 400x (201 Normal, 495 Cancerous)

### 2.3. Data Pre-processing

Image pre-processing is an essential step in any computer vision task and involves operations to give the data a format suitable for training. Deep learning models are computationally expensive and require all input images to have the same shape. The dataset used in this research work contains high-dimensional images with different shapes. We reduced the size of images by reshaping them to the same dimension of 300*×*300*×*3. Moreover, the images were normalized by rescaling the pixel values between 0 and 1. Normalizing input images removes the difference in magnitude between different pixels, which benefits learning [20].

### 2.4. Data Augmentation

The images in the dataset were deficient and had imbalanced classes. DL models, if trained with a small dataset, results in over-fitting, and as a result, the models’ generalization capability becomes very poor. In the case of imbalanced data, the models have poor predictive performance, specifically for the minority class. We overcame these issues by using data augmentation techniques. The images in the minority class (non-cancerous) were augmented by two folds applying geometrical transformations such as horizontal-flip and vertical-flip (Figure 3). Since the pathologists can easily interpret the images from different angles, the flipped images are invariant [21]. Similarly, the overall size of the data was increased by augmenting the images during training.

**Figure 3:**
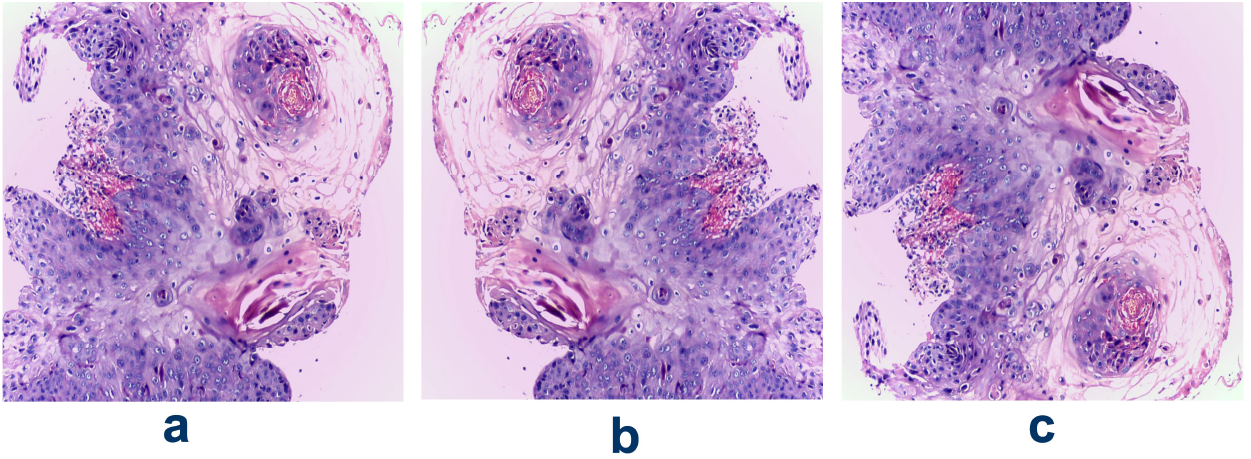
(a) Original image (b) Horizontal-flip (c) Vertical-flip

### 2.5. Network Architectures

#### 2.5.1. VGG16

VGG16 is a sixteen layers deep CNN. It consists of thirteen convolution layers arranged into five blocks, each followed by a pooling operation. The network uses filters of size 3*×*3 for convolution and 2*×*2 size windows for pooling operation. The convolutional stack is followed by two fully connected layers, each consisting of 4096 nodes. The final layer is a softmax layer that assigns a class to each image [22]. The architecture of VGG16 is depicted in Figure 4.

**Figure 4:**
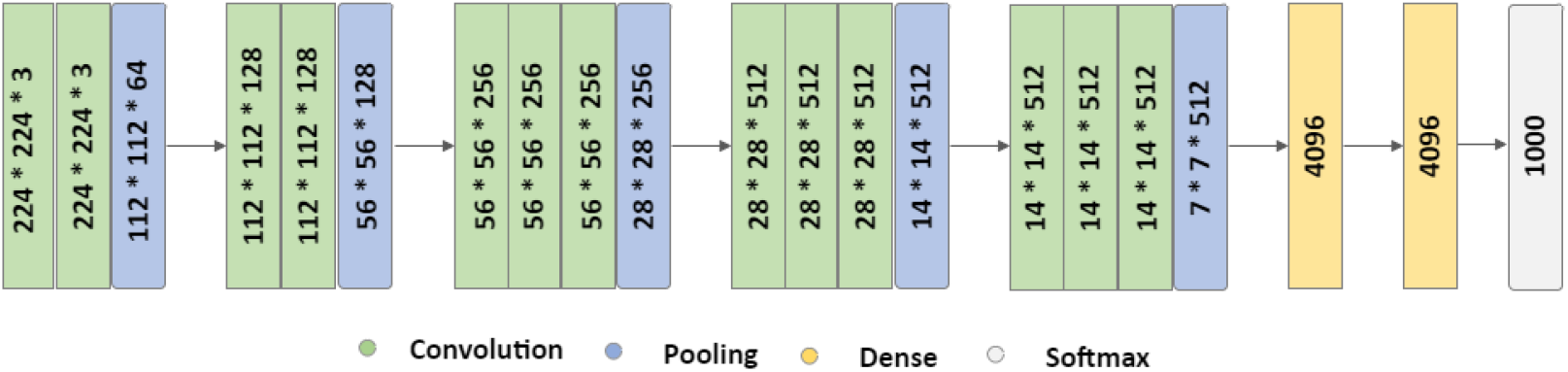
VGG16 architecture consisting of five convolutional blocks (each followed by a pooling operation), two FC layers (each with 4096 nodes) and one Softmax layer (1000 nodes that equals the number of classes in ImageNet dataset) at the end.

#### 2.5.2. InceptionV3

InceptionV3, developed by Christian *et al* [23], is the third version of Google’s Inception CNN. It focuses on reducing the computational cost and the information loss that occurs due to a drastic reduction in input dimension in deeper networks. An important technique used in InceptionV3 to improve computational speed is smaller and factorized convolution filters. For example, the 5*×*5 filters are replaced with two 3*×*3 filters, further factorized into 1*×*3 and 3*×*1 filters. Similarly, the filter banks are widened instead of making deeper to deal with the representational bottleneck. For this purpose, each inception module in the network deploys filters of different dimensions and concatenates their outputs together. Figure 5 shows a module of the Inception network.

**Figure 5:**
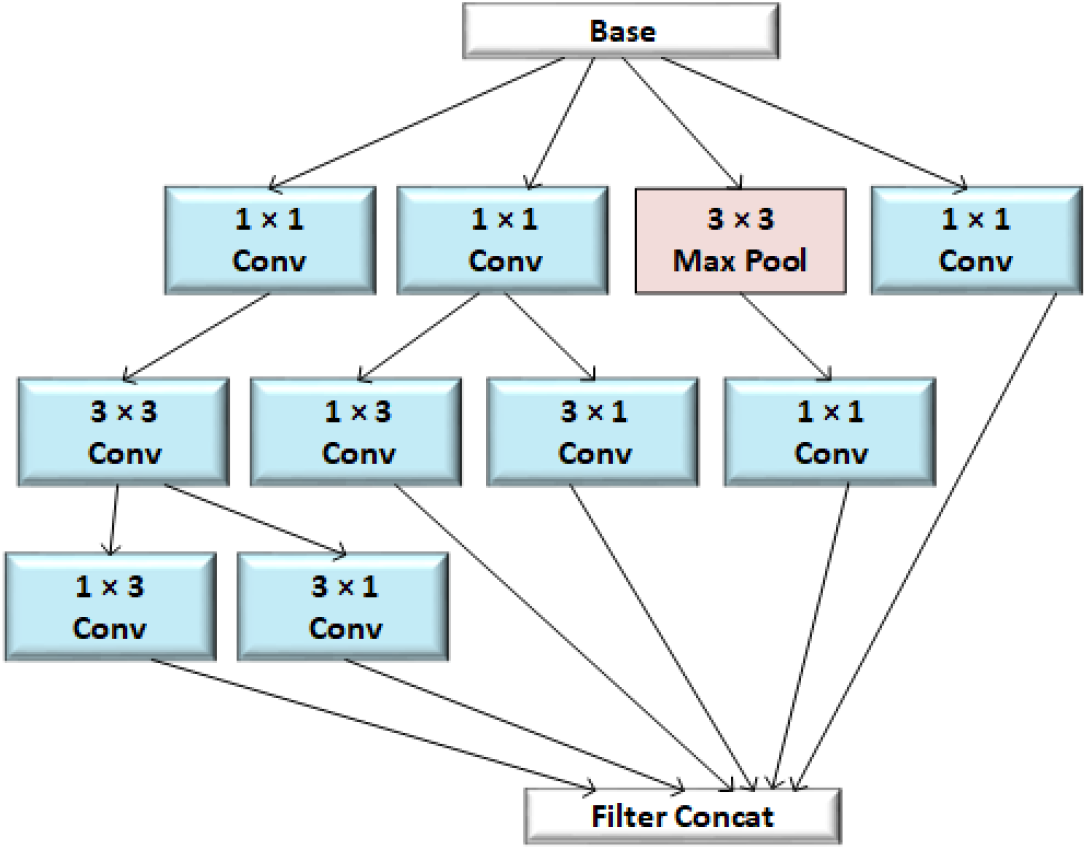
A schematic of an inception module that uses smaller and factorized convolutions and widens the filter bank. 5*×*5 convolutions are replaced with two 3*×*3 filters which are further factorized into 1*×*3 and 3*×*1 filters.

#### 2.5.3. ResNet50

ResNet, which is short for Residual network, is almost similar to a conventional DL model with convolution layers stacked one over the other. The only difference, which makes it a residual network, is the identity connection between the layers. These connections reduce vanishing gradient in deeper architectures and ensure that the higher layers perform at least as well as the lower layers and not worse. Figure 6 depicts an example of a residual block. ResNet50, as the name indicates, is a variant of Residual Networks consisting of fifty layers — forty-eight convolution, one max-pooling, and one average pooling [24].

**Figure 6:**
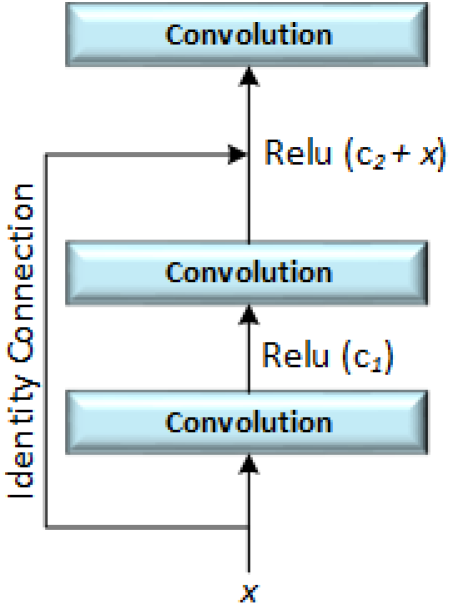
A residual block with an identity connection that passes input(x) directly to a later convolutional layer to minimize vanishing gradient.

### 2.6. Transfer Learning

Transfer learning is a technique in which knowledge learned by a model from one problem is stored and applied to another relevant problem. Using this technique, instead of starting the training process from scratch, the patterns learned by a model previously are adjusted to the new problem. Transfer learning lowers the training time and enables the training of DL models with small data. A standard research practice is to use DL models pre-trained on publicly available large datasets such as ImageNet, CIFAR, etc. These pre-trained models are either fine-tuned or used as feature extractors for the target task (Figure 7).

**Figure 7:**
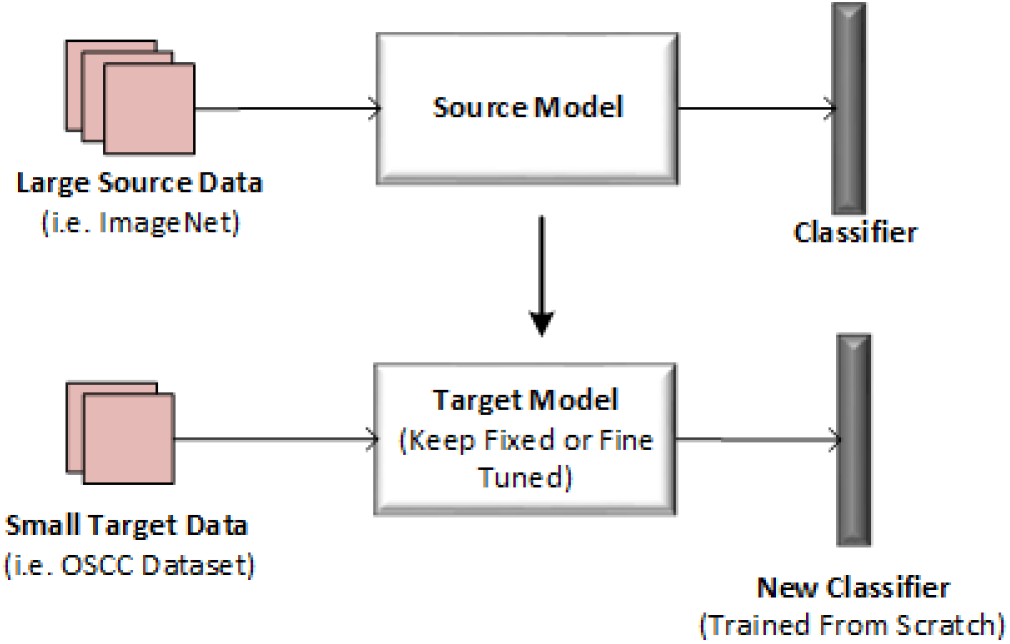
Transfer of knowledge from a source model trained on large data (i.e. ImageNet) to a target model with small data (i.e. OSCC Dataset). The target model is either keep fixed and used as a feature extractor or fine-tuned and adapted to the new task. The newly added classifier to the target model is trained from scratch.

### 2.7. Model Training

We trained our models on 1684 images containing 810 normal and 874 cancerous. VGG16, InceptionV3, and ResNet50 architectures were already trained on a large publicly available dataset called ImageNet (consisting of millions of images with 1000 classes). To train these models for our OSCC detection task, we utilized the concept of transfer learning in two different ways.

#### 2.7.1. Fine Tuning Individual Models

We replaced the actual classifier (softmax layer with 1000 nodes) in each pre-trained model with a new one (sigmoid layer with 1 node) for binary classification of OSCC images. During training, for each model, the bottom layers were kept fixed and not retrained, while a few top layers and the appended classifier were fine-tuned (figure 8 (a)). The primary aim of fine-tuning the upper layers was to adapt the formerly learned advanced patterns from the ImageNet to our OSCC detection task.

**Figure 8:**
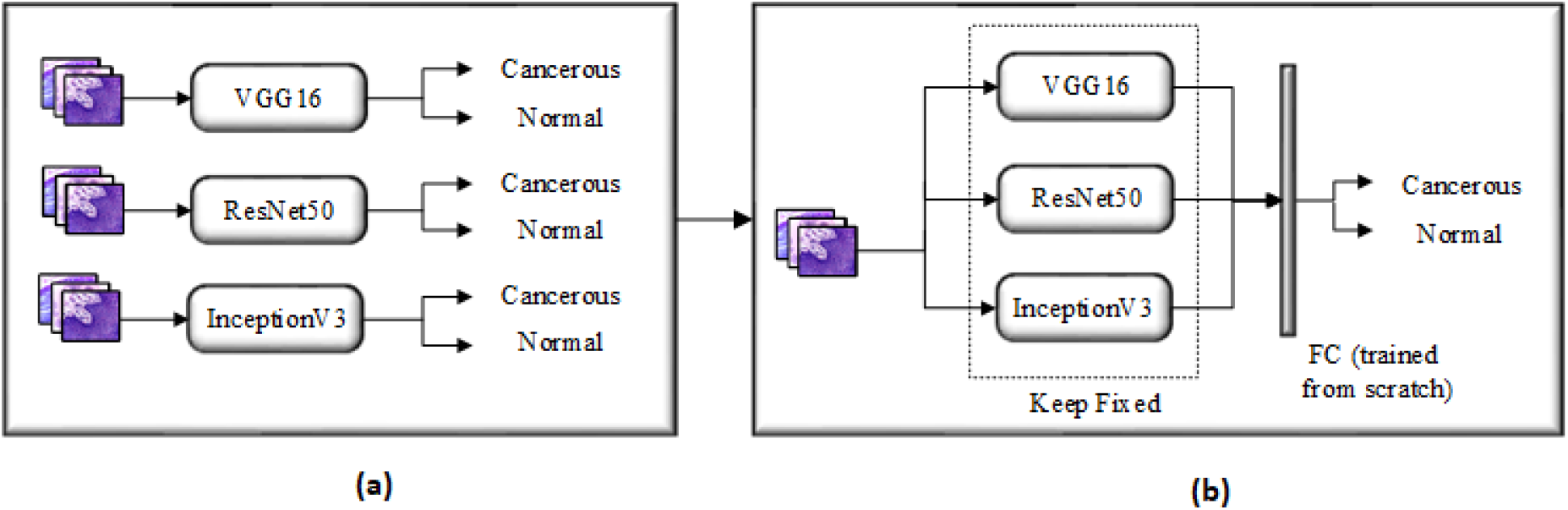
Flow diagram of the model training (a) Pre-trained VGG16, ResNet50 and InceptionV3 are finetuned separately on the OSCC dataset (b) Features extracted by fine-tuned models are concatenated and passed through an FC layer. The concatenated model is not retrained while the FC layer is trained from scratch.

#### 2.7.2. Concatenated Model as Feature Extractor

We designed a new model by concatenating the features extracted by the three fine-tuned models. A fully-connected (FC) layer with ten nodes, and a classifier was added to the model at the end. In this case, the three models were kept fixed and used as feature extractors during the training. The newly added FC layer and classifier were trained from scratch to help the model learn more valuable information from the combined features 8 (b)).

### 2.8. Functions and Parameters

We used Sigmoid as a classification function, Binary cross-entropy as loss function, and Adam as an optimizer for each model. The learning rate was set to 0.001 for individual models while 0.0001 for the concatenated model. As the concatenated model had fewer trainable parameters than the individual models, it used a large batch size and fewer epochs (converged quickly). Similarly, different augmentations were applied to the training data on the fly during the training to avoid over-fitting. Table 1 presents various parameters and functions used during the training.

**Table 1:** Functions and parameters used for each model during the training. All the model used the same classification function, loss function, optimizer and geometrical transformations for data augmentation. As the concatenated model has less trainable parameters and is computationally inexpensive, it used less epochs and large batch size for training.

| Function/Parameter | VGG16 | InceptionV3 | ResNet50 | Concatenated |
| --- | --- | --- | --- | --- |
| Classification Function | Sigmoid | Sigmoid | Sigmoid | Sigmoid |
| Loss Function | Adam | Adam | Adam | Adam |
| Optimizer | Binary-cross Entropy | Binary-cross Entropy | Binary-cross Entropy | Binary-cross Entropy |
| Epochs | 50 | 50 | 50 | 10 |
| Batch Size | 32 | 32 | 32 | 64 |
| Learning Rate | 0.001 | 0.001 | 0.001 | 0.0001 |
| Horizontal-Shift | 5% | 5% | 5% | 5% |
| Vertical-shift | 5% | 5% | 5% | 5% |
| Zoom | 5% | 5% | 5% | 5% |
| Rotation | $40^0$ | $40^0$ | $40^0$ | $40^0$ |

### 2.9. Models Evaluation

We validated the performance of models with 120 images, including an equal number of instances from both classes. The test dataset contained images with both 100x and 400x magnification. Moreover, The train/test split was performed before the augmentation, which means the test dataset contained original images.

To evaluate the performance, we calculated accuracy, precision, Recall, F1-score, specificity, and AUC value for each model. These statistical metrics are based on True Positives (TP), False Negatives (FN), False Positives (FP), and True Negatives (TN). Here, TP and TN represent the number of correctly identified cancerous and normal images, while FP and FN denote misclassified normal and cancerous images, respectively.

The accuracy scores tell how often the models produced correct result (Eq. 1).

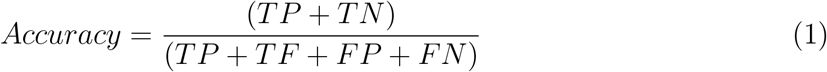

Precision score determines the ratio of correctly identified cancerous images to all the images predicted by a model as cancerous (Eq. 2). In other words, precision reflects a model’s consistency with respect to cancerous outcomes.

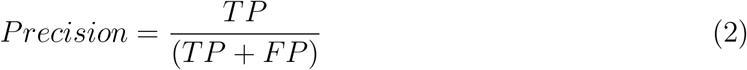

Recall calculates the ratio of correctly identified cancerous images to all the cancerous images in the test data (Eq. 3).

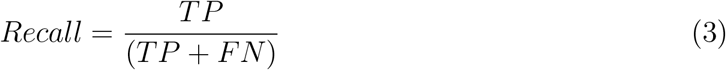

Specificity performs the same operation as recall but for normal images (Eq. 4).

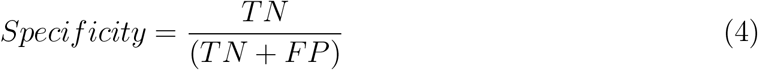

F1 score represents a weighted average of precision and recall (Eq. 5).

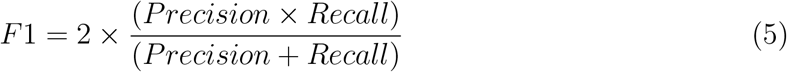

Receiver Operating Characteristic (ROC) plots TP rate (Eq. 6) versus FP rate (Eq. 6) and helps us understand the relationship between correctly classified cancerous and misclassified normal images at different thresholds. Area Under Curve (AUC) is a scalar value ranging between 0 and 1 and represents how well our models discriminated between normal and cancerous images.

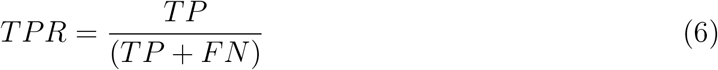

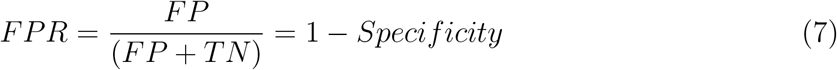

## 3. Results and Discussion

Confusion matrices in figure 9 show the performance of each model on the test data. It can be observed that the concatenated model, where all of the three models were used in combination, has the highest TP and TN and lowest FP and FN values. Among the three models used individually, the InceptionV3 model achieved better performance than ResNet50 and VGG16. This is also obvious from table 2 where the concatenated model achieved the highest accuracy, precision, recall, F1-score, and specificity values (96.66%, 95.16%, 98.33%, 96.71%, and 95.00%) followed by InceptionV3 (94.16%, 93.44%, 95.00%, 96.27%, and 93.33%), ResNet50 (90.83%, 88.88%, 93.33%, 91.05%, and 88.33%), and VGG16 (89.16%, 87.30%, 91.66%, 89.42%, and 86.66%). Figure 10 shows a plot of ROC-AUC curves where each color represents a different model. The highest and lowest AUC values were achieved by the concatenated model (0.997) and VGG16 (0.951), respectively.

**Table 2:** Results for the four network architectures

| Model | Accuracy (%) | Precision (%) | Recall (%) | F1-score (%) | Specificity (%) |
| --- | --- | --- | --- | --- | --- |
| VGG16 | 89.16 | 87.30 | 91.66 | 89.42 | 86.66 |
| InceptionV3 | 94.16 | 93.44 | 95.00 | 96.27 | 93.33 |
| ResNet50 | 90.83 | 88.88 | 93.33 | 91.05 | 88.33 |
| Concatenated Model | 96.66 | 95.16 | 98.33 | 96.71 | 95.00 |

**Figure 9:**
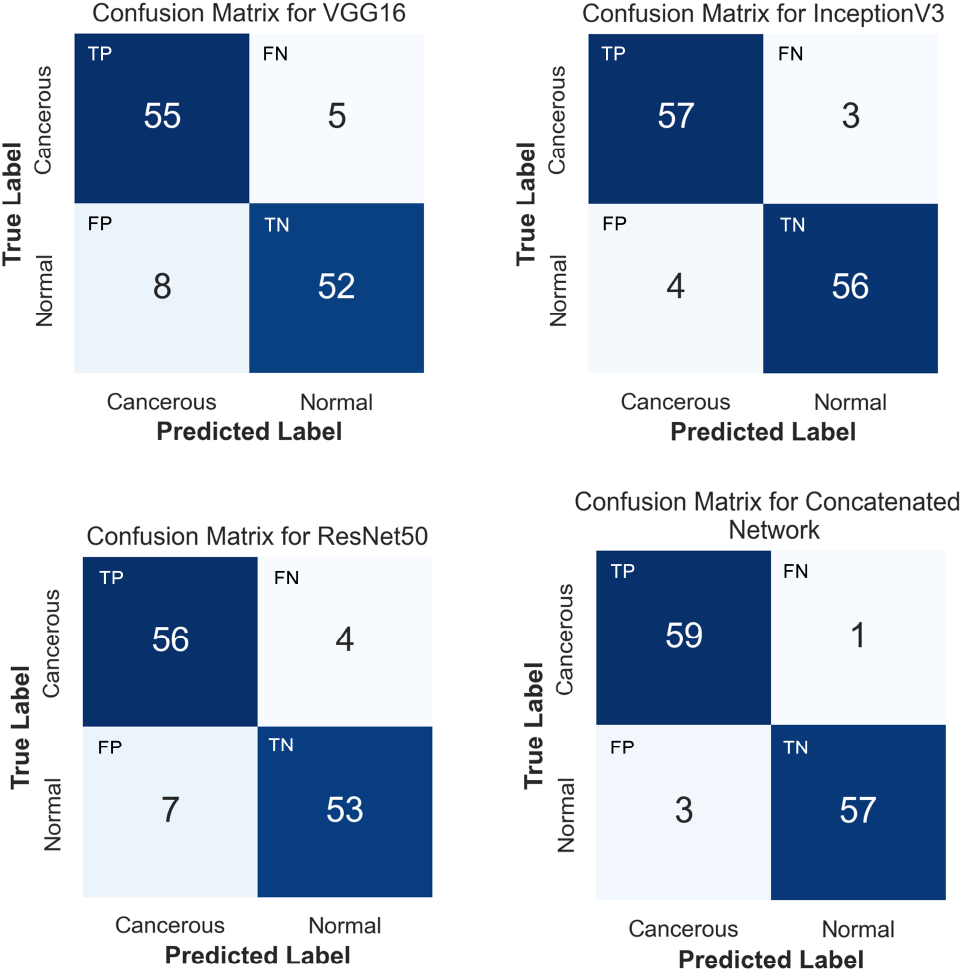
Confusion matrices for the network architectures

**Figure 10:**
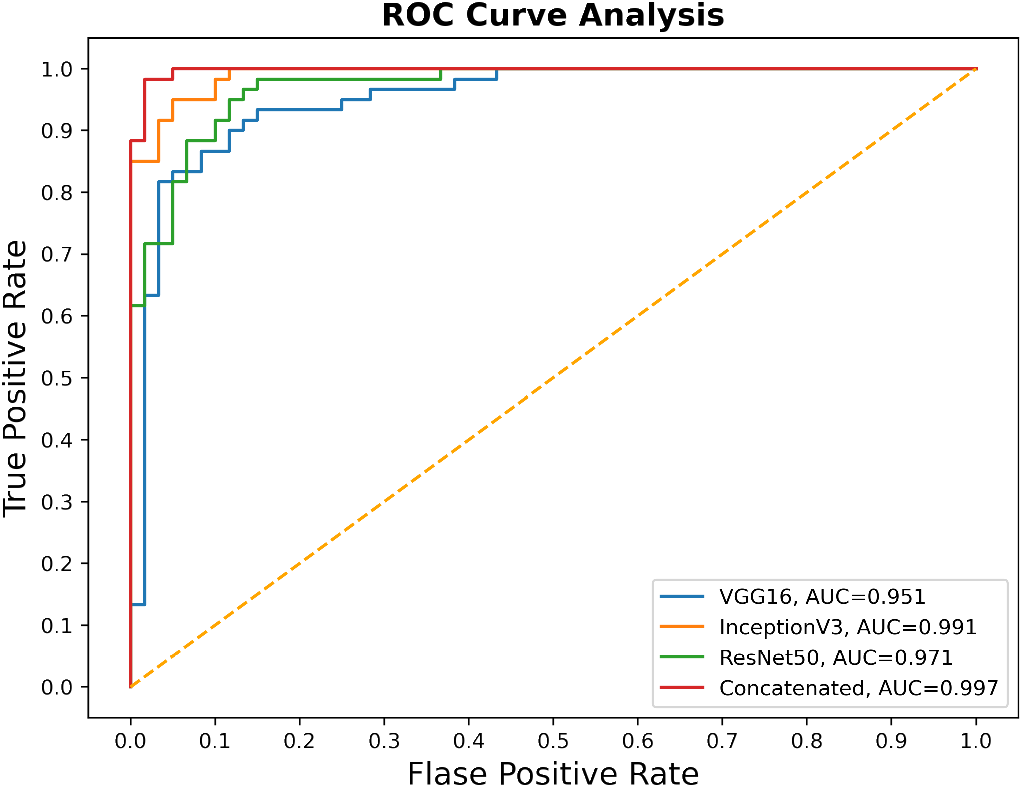
Plot depicting ROC curve and AUC value for each model

It is understood from the confusion matrices (figure 9) and table 2 that the concatenated model performed better in detecting OSCC and not detecting false cases and outputs the best overall accuracy. Individual models, particularly VGG16 and ResNet50, performed much better on identifying cancerous images than normal, which is evident from their high FP and low specificity and precision values. This is because we had imbalanced data with poor representation of the normal class. Interestingly, the concatenated model improved the performance not only for identifying cancerous images but also normal images (lowest FP and FN). Similarly, a high AUC value of 0.997 indicates that the concatenated model has the excellent capability of differentiating between the two classes.

A recent study by Rutwik *et al* also performed the classification of OSCC images into normal and cancerous using different pre-trained DL models. The highest accuracy of 91.13% was obtained by ResNet [18]. Our concatenated model significantly improved the classification of OSCC images by achieving an accuracy of 96.66%. The usefulness of combining features from multiple models has also been reported for the detection of breast cancer [21]and covid19 [25]. Sanaullah *et al* reported that the high performance of concatenated models is because the combined features may contain multiple patterns, i.e., circularity, roundness, compactness, etc., from a single descriptor [21]. This is the reason that in our study, the combination of multiple pre-trained CNN architecture boosted up the performance of transfer learning and may replace the use of traditional single model CNN architecture.

## 4. Conclusion

In this paper, we performed classification of oral histopathological images into normal and cancerous classes using three different pre-trained DL architectures (VGG16, InceptionV3, and ResNet50). First, the models were fine-tuned individually and used for the classification task. The fine-tuned models were then combined by concatenating their extracted features. Finally, we compared the performance of the three fine-tuned models with the concatenated model. It was observed that the concatenated model yielded the best results and outperformed the individual models.

A small test data may not completely represent the data collected in the real world and may not have good coverage of the distribution in training data. The OSCC dataset used in this study was both small and imbalanced with 290 normal and 934 cancerous cases. We could use only 120 images for evaluating the model, which may be acceptable, but not ideal for translating to real-world applications. In future, we aim to improve this by generating large datasets, that ideally have matched classes. Similarly, we may also consider using different cross validation methods that are more appropriate for validating the model on a large data.

## Data Availability

We used a publicly available dataset, published by Tabassum et al, Data in Brief, 2020.

